# A Quantitative Evaluation of COVID-19 Epidemiological Models

**DOI:** 10.1101/2021.02.06.21251276

**Authors:** Osman N. Yogurtcu, Marisabel Rodriguez Messan, Richard C. Gerkin, Artur A. Belov, Hong Yang, Richard A Forshee, Carson C. Chow

## Abstract

Quantifying how accurate epidemiological models of COVID-19 forecast the number of future cases and deaths can help frame how to incorporate mathematical models to inform public health decisions. Here we analyze and score the predictive ability of publicly available COVID-19 epidemiological models on the COVID-19 Forecast Hub. Our score uses the posted forecast cumulative distributions to compute the log-likelihood for held-out COVID-19 positive cases and deaths. Scores are updated continuously as new data become available, and model performance is tracked over time. We use model scores to construct ensemble models based on past performance. Our publicly available quantitative framework may aid in improving modeling frameworks, and assist policy makers in selecting modeling paradigms to balance the delicate trade-offs between the economy and public health.

## Introduction

Epidemiological models of COVID-19 have proliferated quickly but it is unclear how well they forecast true values of key underlying variables: the number of infected and dead individuals. Establishing the quality of inference and predictive power is essential to make data-driven public health decisions, including those that manage the delicate trade-offs between the economy and public health. Model evaluation is therefore of critical importance (1, 2).

We have created a uniform objective scoring system for COVID-19 models that assess their predictive performance. A scoring system has previously been deployed to assess Flu prediction models in the FLUSIGHT contest two years ago (3, 4). For COVID-19, investigators have been collating COVID-19 model epidemiological forecasts from multiple research groups in the COVID-19 Forecast Hub (covid19forecasthub.org). These results are concurrently presented on the CDC coronavirus forecasting website. While this effort provides insight into various model forecasts and their various assumptions, it stops short of providing an actual score for cumulative predictive performance.

There have been efforts to assess the accuracy of published and unpublished COVID-19 models. Some have focused on evaluating model performance on their weekly cumulative predictions. For instance, two different studies (5, 6) assessed the performance of different models with the median absolute percent error (MAPE) of cumulative deaths. Fried-man et al. (5), observed that the calculated MAPE increased for longer forecasts, and the best performance model varied by region. Others have focused on evaluating model performance based on weekly incident case forecasts by ranking them according to the mean weekly percentile (7), while Leaderboard (8) uses the root mean squared error of weekly new deaths and reporting recent and running average performance of eight models. Another group evaluated ensemble models strictly containing probabilistic forecasts by computing a weighted interval score (9). They found that combining forecasts in various ways consistently leads to improved performance over single model forecasts.

Here, we propose to score individual models using a Leave-Forward-Out-Cross-Validation scheme. The score is computed by taking the log of the likelihood of the model forecasts on current data using the model predictions from the past. Each weekly projected quantity is scored separately, making it possible to assess model accuracy by the forecasted time increment. Thus, we have a matrix of scores where each entry is the computed log-likelihood for a model fitted to observed data up to a certain date of a quantity for each week forward from that date. This matrix continually expands in size as new weekly forecasts are made and new forecasting teams join the COVID-19 Forecast Hub collaborative. A global score is computed from the matrix by averaging over the desired elements. Some models may do well for short time predictions but not for longer ones and vice versa. Our score keeps track of how models improve (or degrade) over time and how far into the future they can reliably forecast. Our score dashboard is available at www.covidforeca.st.

## Methods

### Obtaining COVID-19 Forecast Hub Data

Forecasts of models are stored in the data-processed folder of the COVID-19 Forecast Hub repository at github.com/reichlab/covid19-forecast-hub. The repository contains forecasts of weekly cumulative and incidental death counts, weekly incidental COVID-19 positive case counts and daily incidental hospitalizations for the whole US and for the individual states. Each forecast includes a ‘target end date’, *t*, which is the date when ground truth *G*_*t*_ is observed and a ‘forecast date’, *d*, which is the date when the forecast was made. The forecast submissions include Forecast Hub standard quantiles for the forecasted distributions. Death count forecasts need to contain 23 standard quantiles (i.e. c(0.01, 0.025, seq(0.05, 0.95, by = 0.05), 0.975, 0.99)) while weekly incidental case counts require 7 (i.e. c(0.025, 0.100, 0.250, 0.500, 0.750, 0.900, 0.975)). We only evaluate forecast entries that obey these guidelines. We focused on weekly cumulative death counts and weekly incidental case counts for the US (Fig. S1). Ground truth data (*G*) for case counts, deaths, and hospitalizations are also available in the Forecast Hub repository through JHU CSSE Group (10) and HealthData.gov. For week-ahead forecasts, a week runs from Sunday through Saturday by definition, and we download the Forecast Hub data every Sunday evening and update the model scores. Sometimes a modeling team deploys multiple forecast entries on different dates targeting the same future target end date and in such cases, we use only the forecast with the latest forecast date.

### Calculation of model forecast scores

To calculate scores of epidemiological forecasts on the COVID-19 Forecast Hub, we first converted the reported quantiles into approximate probability density functions (PDFs) and then computed the log-likelihood of the held-out future observation as depicted in Figure 1.

**Fig. 1.**
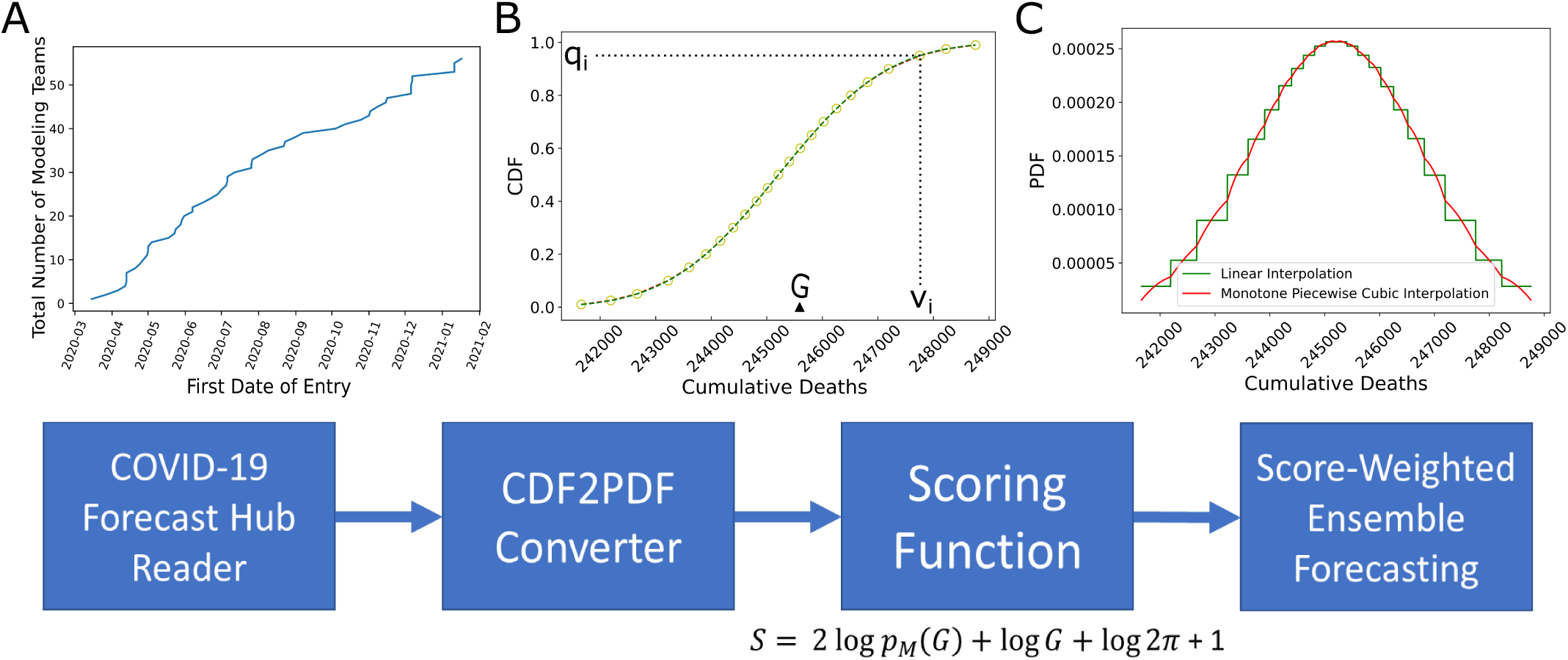
Scoring framework and analysis. A. Total number of teams which deployed US country-level epidemiological forecasts on COVID-19 Forecast Hub as of January 24, 2021. B. Scoring starts with reading forecast data available at COVID-19 Forecast Hub. An example forecast is shown for the model BPagano:RtDriven forecast made on 2020-11-9 targeting cumulative number of deaths on target end date 2020-11-14 (as denoted by *G*). Each forecast has a set of quantiles *q* and a set of corresponding values *v*. C. We calculate probability density functions using forecast data {*q,v*}(details in Methods). We apply our scoring function on every forecast available. The past performances of models can be used to form score-weighted ensemble model forecasts.

### Computation of model likelihood

The model predictions for a model, *M*_*d,t*_, are provided as standard quantiles **q** of the forecasted cumulative distributions for the case and death counts, **v**, computed using data up to a forecast date, *d*, for a prediction at a target end date, *t*. We convert this to a model likelihood by computing the PDF and evaluating at the corresponding ground truth value, *G*_*t*_. From the quantile data, we upsample **v** to create a regularized one dimensional grid **V** such that **V** = [floor(*v*_min_)+0.5 : ceil(*v*_max_)-0.5] and thus each grid point is separated by a distance Δ*V* = 1. This Δ*V* is held constant across all models. Next, we calculate **Q** = *F* (*V*_*i*_), the cumulative probabilities corresponding to **V**, based on Piecewise Cubic Hermite Interpolating Polynomial (PCHIP) interpolation of **q**. Using Python NumPy gradient function we find the derivative of **V**, *F*^*t*^(*V*) = *f* (*V*), which is the approximate PDF. Finally, we find the grid point interval [*V*_*i*_, *V*_*i*+1_] in **V** containing the true value, *G*_*t*_ (an integer). If both grid points of the PDF are available, then we define the probability of having *G*_*t*_ given the model as 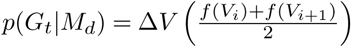. Otherwise, if the actual data falls outside of the forecast prediction interval, we assign *p*(*G*_*t*_ |*M*_*d*_) = 0 (even though this probability is non-zero for a distribution on semi-infinite support, it is insignificantly small). We validated our algorithm by finding small error between the integrated *p*(*G*_*t*_ |*M*_*d*_) values and the forecaster provided maximum quantile ranges (Fig. S2).

### Scoring function

We constructed a scoring function that rewarded models for assigning high probabilities to the true values. Thus, models with broad predictive distributions are penalized compared to models with narrower distributions even if the true values are at the mode in both distributions. Conversely, models with narrow distributions are severely penalized if the true value falls outside of their predicted distribution.

To this end, we start with the log-likelihood for a model *M*_*d,t*_ given the held-out forecast target *G*_*t*_. However, the problem with the log-likelihood alone is that the score will vary depending on the distribution of *G*_*t*_, which can vary over time. To alleviate this problem, we normalize the score by subtracting away an “optimal log-likelihood” so that

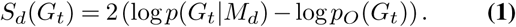

This is similar to the negative of the deviance used for generalized linear models. In place of a saturated model prediction, we assume a normal likelihood (for large rates a Poisson distribution is near normal), then we can write

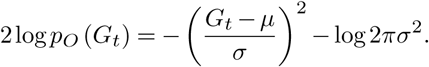

with an expectation of

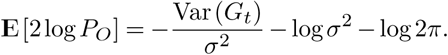

The maximum is given by *σ*^2^ = Var(*G*_*t*_), and setting Var(*G*_*t*_) = *G*_*t*_, as expected for a Poisson distribution (although the actual data is often overdispersed) we define our score as

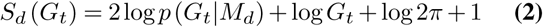

We note that *S*_*d*_(*G*_*t*_) is−∞ if *G*_*t*_ falls outside of the forecast prediction interval of a model *M* at any {*t, d*}pair, since we assign *p*(*G*|*M*) = 0 in such cases.

### Overall model performance evaluation

Model scores depend on many intrinsic (e.g. changes to model assumptions) and extrinsic (e.g. epidemiology) factors, and most importantly on time. Thus, an overall model performance evaluation spanning through the entirety of available data of the pandemic can provide insight into all of these components (albeit in aggregate). To make fair comparisons, we analyze groups of models whose forecasts have the same target time horizon (e.g. 1 week ahead or 6 weeks ahead) together and have separate leader boards for each time horizon. We evaluate a model’s performance on date *t* using two metrics: (1) median past score *MS* and (2) mean past model score performance ranking among other comparable models *R*_*t*_ such that

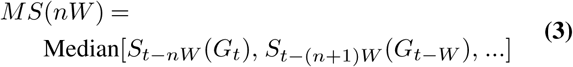

and

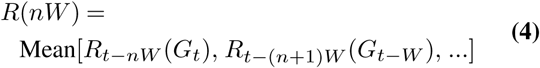

where *nW* is the n-Week forecast horizon. The leader board considers the time frame starting from July 4, 2020. We do not include models with number of forecasts less than 50% of the number of possible weeks in the time frame in the leader boards (Fig. S7).

Additionally, we used Median Absolute Deviation (MAD) to measure score variability of the models over time. For each forecast type (case or death) and each forecast horizon, we have a set of scores for a model’s forecasts from July 4, 2020 onward. We find the median of the set. Then, we find the absolute difference between the elements of the set and the calculated median. This results in a set of absolute deviations. We report the median of that set for each forecast type and each forecast horizon.

### Score-weighted Ensemble Formation

We generate the ensemble model forecast values **v**_**e**_ corresponding to the standard set of quantiles **q** by using weights informed by past model scores (Fig. S4). Assuming there are *n* models available at a given forecast date *d* with a particular target end date *t*, to obtain **v**_**e**_, we weigh the constituent model forecast values **v**_**M**_ corresponding to the *i*^*th*^ quantile *q*_*i*_ (see (11))

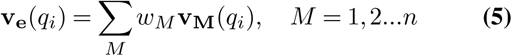

where *w*_*M*_ is the normalized weight of each model. There are many ways of formulating model weights in an ensemble, but we used median of exponentiated past scores of a model up to the date of forecast *d* as weight such that

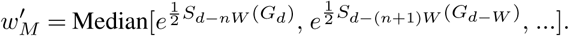

We normalize 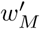 over all available model weights to calculate *w*_*M*_.

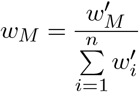

Hence, an unweighted ensemble has all *w*_*M*_ equal to 1*/n*. Also, we note that when forming time horizon-specific ensembles (e.g. 4-week ahead ensemble), we use scores and averages of that particular time-horizon only and do not involve others (e.g. 1-week ahead or 6-week ahead). We call our score-weighted ensemble model Sweight with the modeler team name FDANIHASU.

### Development of the Web-based User-Interface

All model forecasts (including those of the ensemble described above) are scored using Equation 2. We have developed leader boards based on running median scores and running average rankings of the models. We have developed a Python Plotly/Dash-based dashboard for the leader board and model score analysis along with various plots. The dashboard is updated every Sunday of the week and is available at the website www.covidforeca.st.

### Systematic review of model types

To assess model performance based on similarity between model types and assumptions, we performed a meta-analysis of the models available on the COVID-19 Forecast Hub. As of January 24, 2021, there were 90 models deployed, and we reviewed 63 of those models which reported whole US population forecasts (weekly cases and cumulative deaths) along with the required pre-designated quantiles to perform scoring. In order to review each model, we first classify them under: *deterministic, stochastic* or *statistical*. A model was classified as *deterministic* if it is built using macro-level compartments representing a group of individuals and the dynamics are defined using average transition rates between compartments. A *stochastic* model is built at micro-level states occupied by discrete individual persons, randomness is introduced, and the transitions are defined with probabilities. A model was classified as *statistical* if statistical techniques were applied to analyze a data set and regression-type models were utilized. Deterministic models were categorized further into compartmental and metapopulation. Compartmental models assume a continuous population that is divided into a number of states (e.g. susceptible, infected, and recovered) (12). Metapopulation models are usually in the form of connected compartment models of subpopulations defined in terms of geographical regions and incorporating random mixing within these sub-populations (12), therefore adding complexity to compartmental types.

## Results

### COVID-19 Forecast Hub data characterization

The number of models on the COVID-19 Forecast Hub increased substantially since March 2020 (Figure 1). We focused on weekly incidental case forecasts and cumulative death counts (Fig. S1). We identified 39 models for the weekly case counts and 54 models for cumulative death counts. The majority of the forecasts target less than 4-week ahead COVID-19 epidemiology. There are 7402 unique entries for cumulative death count forecasts and 3759 for the weekly incidental case counts as of January 24, 2021 and the cumulative death counts forecasts span a much larger time frame (up to 21 weeks ahead). When we look at the distribution of scores, we see that scores decrease for long-term forecasts substantially and models are in general much better (higher score) at forecasting cumulative death counts than in forecasting weekly incidental case counts (Figure 2 panels C and D and Figure S3). We also noticed that the models failed to capture abrupt changes in the observed case counts, specifically first week of July 2020 and mid November 2020 (Figure S3 and 3).

**Fig. 2.**
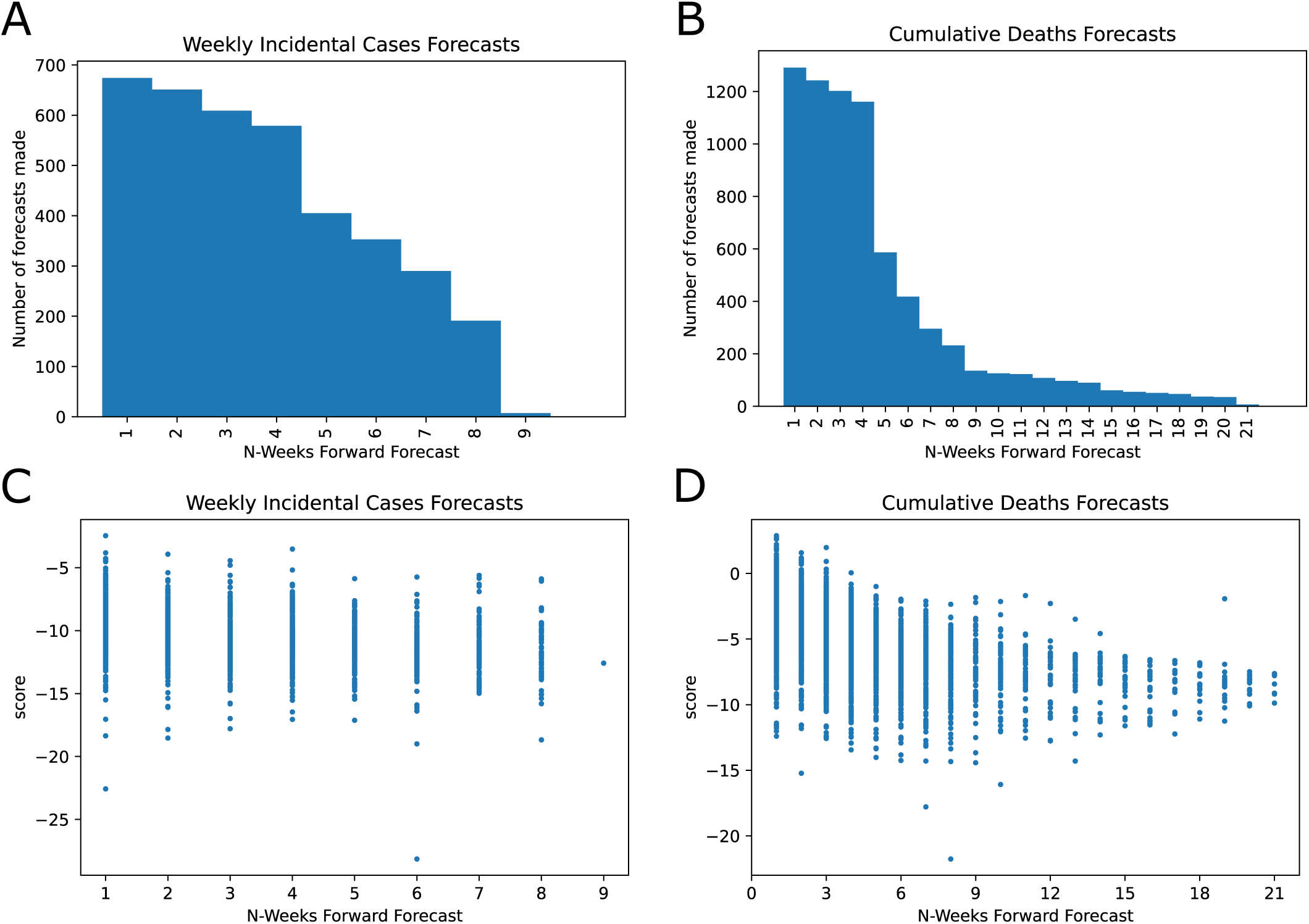
COVID-19 Forecast Hub data general review and scores. A. Histogram of weekly incidental case count forecasts for the US. B. Histogram of cumulative deaths forecasts for the US. C and D shows the scatter plot for all scores as a function of the forecast horizon. C. Weekly incidental case forecast scores. D. Cumulative death count forecasts

**Fig. 3.**
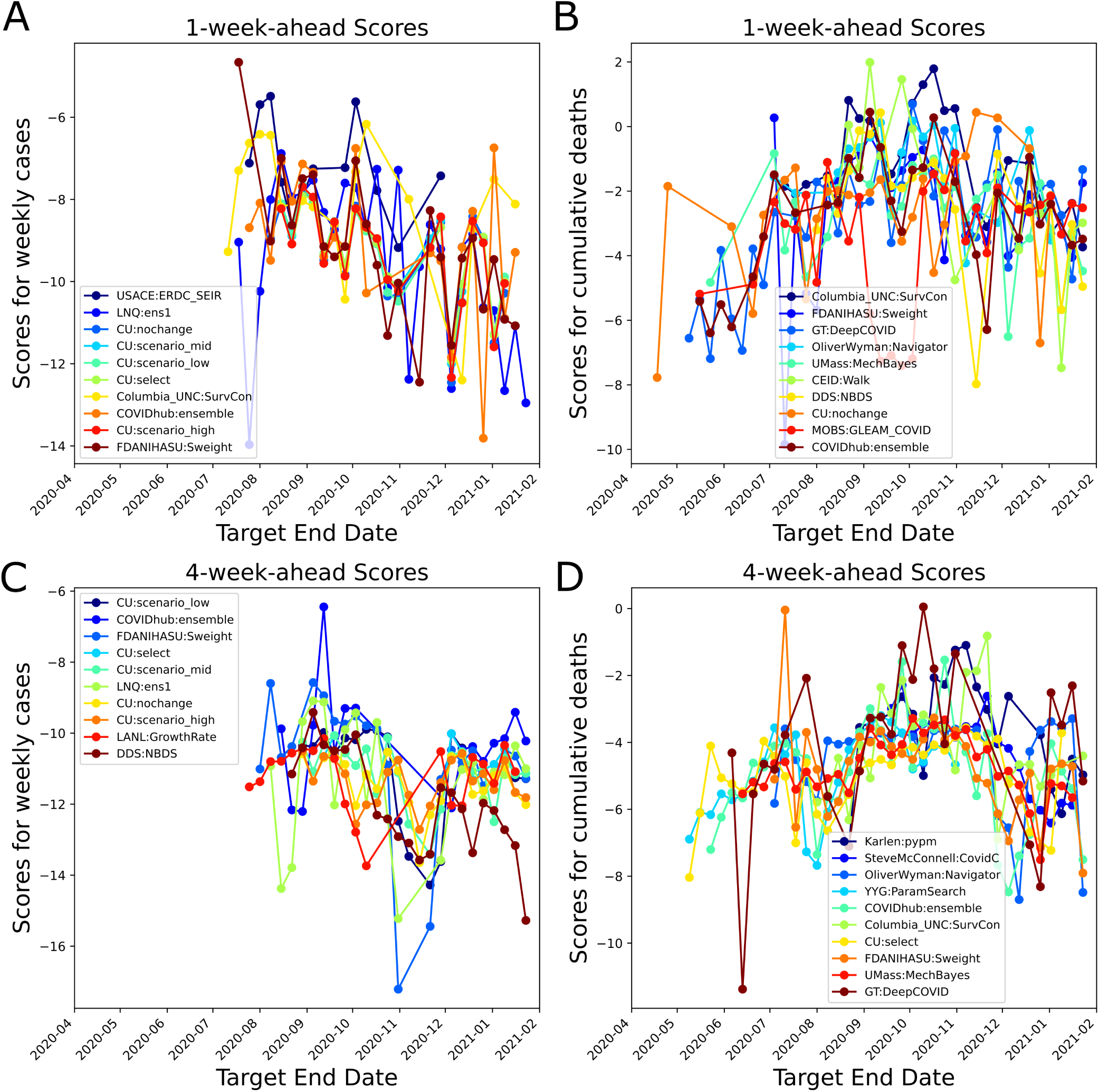
Average 1-week- and 4-week-ahead forecasting performances shown for the top 10 models based on running average scores (Eq. 3) (blue color for the best performing on average and dark red for number 10). FDANIHASU:Sweight model is the score-weighted ensemble presented in this work. A. Average scores for Weekly incidental COVID-19 Cases forecasts (1-week-ahead performance). B. Average scores for cumulative death count forecasts (1-week-ahead performance). C. Average scores for Weekly incidental COVID-19 Cases forecasts (4-week-ahead performance). D. Average scores for cumulative death count forecasts (4-week-ahead performance).

### Score-Weighted Ensemble Forecasting

We formed un-weighted and score-weighted ensemble models which forecast US weekly incidental case counts and US cumulative death counts over the horizon of 1-week to 6-weeks (see Methods). We calculated the scores of the unweighted and weighted ensembles at each target end date. In general the score-weighted ensemble performed better for 1-week ahead forecasts (Fig. 4). In the early phases of the ensemble (i.e. the first two weeks of July 2020) the number of models within the ensemble for cases was small (less than 5). Additionally, the number of past score samples were limited, which reduced the score-based model credibility for the ensemble. However, as we reached December 2020, we see that changes in the epidemiological dynamics did not impact the score-weighted ensemble as much as the unweighted ensemble (see the declines in scores in December 2020, for 4-week-ahead forecasts, 4B). The differences between the weighted and un-weighted ensemble forecast scores are less significant for the weekly incidental case counts (Fig. S6). Moreover, as an alternative quantitative measure of forecast performance of our ensemble model, we calculated the median of the difference between Sweight’s scores and the median of all available scores across the pandemic over different forecast horizons (large positive difference means better ensemble model performance). Based on our calculations, Sweight performed consistently better and had a higher score than the average model for 1 through 6-weeks-ahead horizon (Table 1). We also observed that Sweight ensemble model tends to make more stable forecasts based on the MAD calculations (Fig. S5).

**Table 1.** Performance evaluation of the score-weighted ensemble model (FDANI-HASU:Sweight) forecasts over time for US cumulative death and weekly incidental cases. We compared median difference in scores (Ensemble -Average Model) for different forecast horizons (i.e. 1-through 6-weeks-ahead). Across all horizons and forecast target types the score-weighted ensemble model performed better based on this measure (i.e. positive values).

| Median Difference in Scores (Ensemble - Average Model) |  |  |
| --- | --- | --- |
| Weeks-Ahead | Case Forecasts | Death Forecasts |
| 1 | 0.766683 | 2.084539 |
| 2 | 1.144867 | 1.171649 |
| 3 | 1.612646 | 1.031703 |
| 4 | 1.317739 | 1.180398 |
| 5 | 2.280982 | 2.074885 |
| 6 | 2.034455 | 2.743361 |

**Fig. 4.**
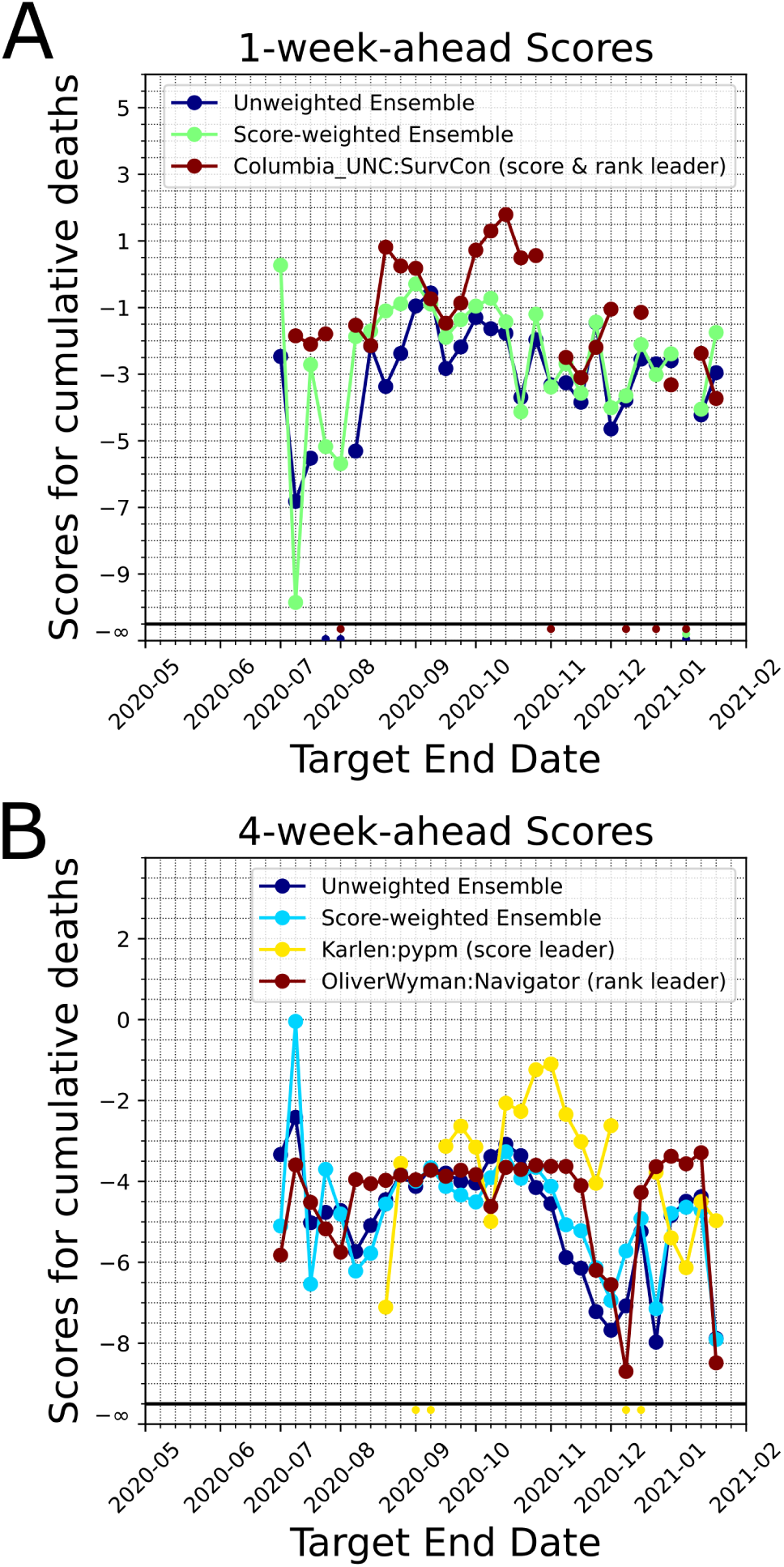
Comparison of scores of unweighted and score-weighted ensemble models for the cumulative death counts. For comparison purposes we plot the leading model’s score (based on past performance as of the last target end date, by ranking and by median scores). Model forecasts that do not encompass the ground truth *G* would have a score −∞ and this scenario is shown at the bottom of the figure panels. A. 1-week-ahead scores. B. 4-week-ahead scores.

### Modeling strategies score comparably despite varying assumptions

Based on our literature review of models, about 54% of the models used the well-known epidemiological SIR model and its variations (i.e. SIR, SEIR, etc.) as the main framework in both deterministic and statistical models. About 43% of the models used statistical and probabilistic approaches, and about 3% of the models used other types such as combination of statistical and logistic growth model or by averaging the predictions of several models (Fig. 5A). Among these classifications, modelers implemented other methods/theories such as spatiotemporal dynamics, seasonality, ground truth reporting periodicities and delays, mobility, and age-structure to increase specificity and complexity. When we analyze the approach the modelers took to drive their main modeling framework, we see that 27% used purely compartmental modeling, 19% used metapopulation approaches to model the spatial/population network interactions, 38% used statistical/stochastic approaches such as deep learning, and about 16% used ensemble forecasting (Fig. 5B). Although model performances change with time, we have not observed dominance in performance of one modeling category over the others (Fig. 6). Our detailed methodological review of models on COVID-19 Forecast Hub can be found in the Supplementary Materials.

**Fig. 5.**
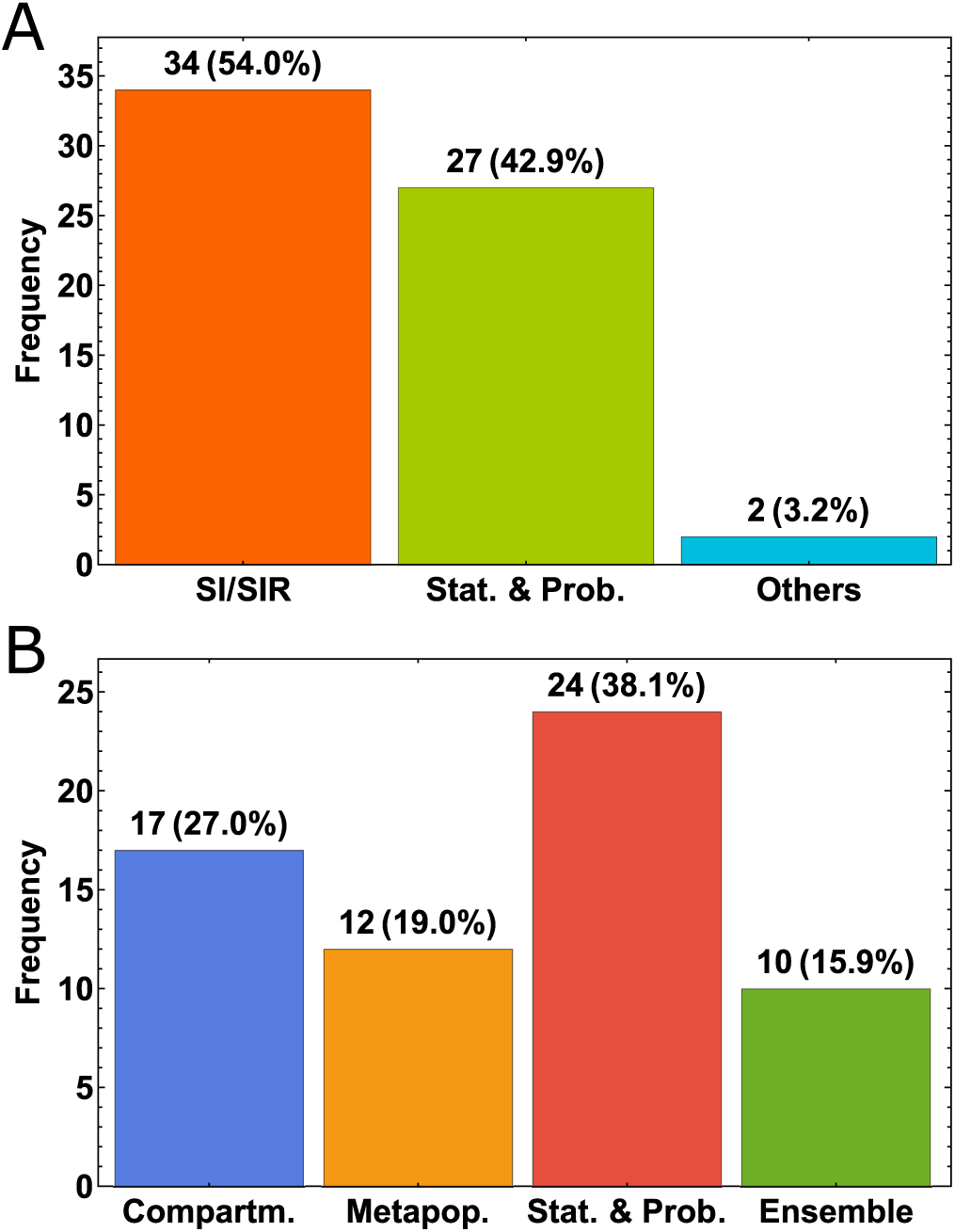
Distribution of the model types. A. General breakdown according to the main framework of the type of models. B. Breakdown by the overarching modeling theme.

**Fig. 6.**
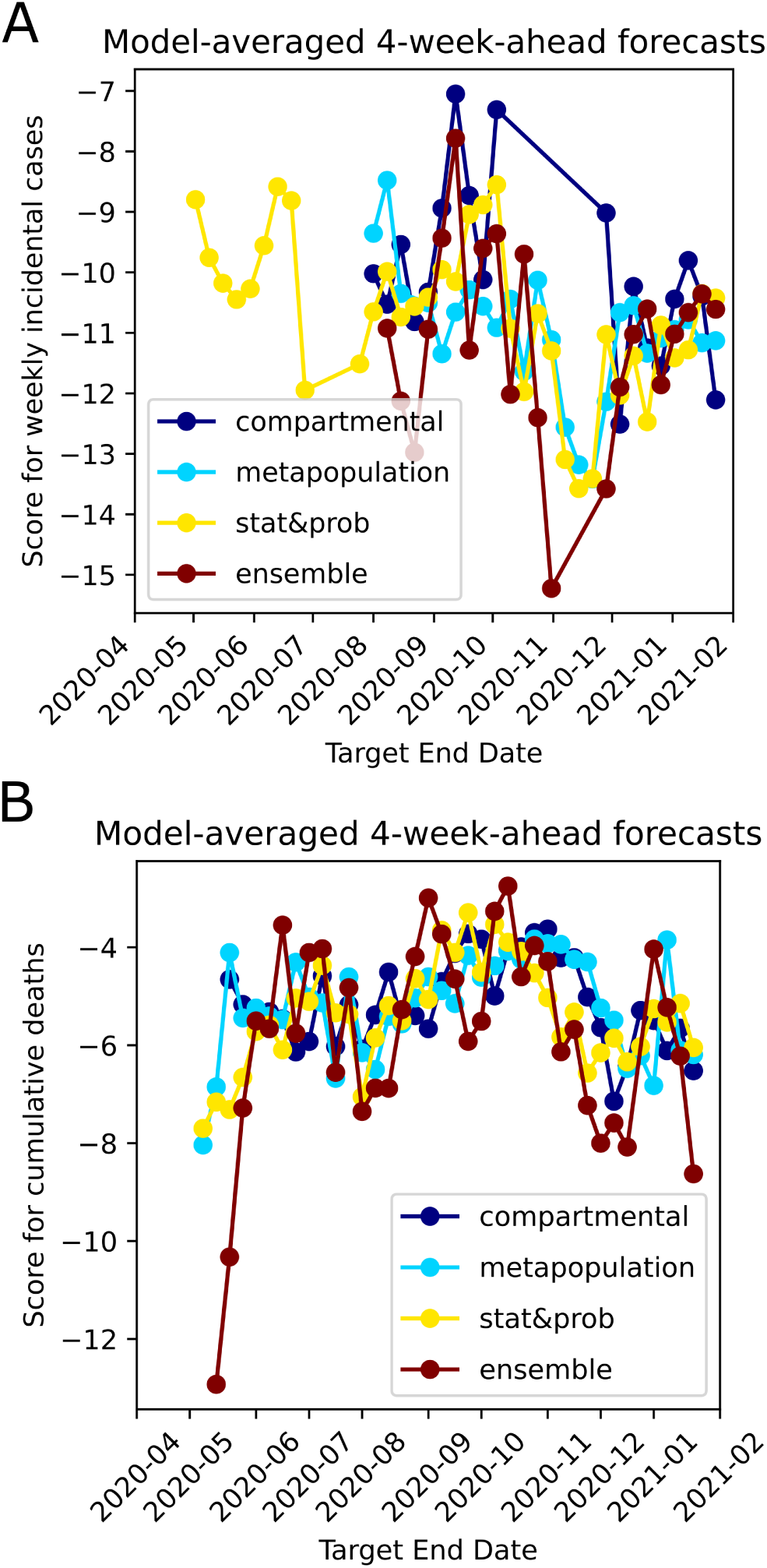
Average 4-week ahead forecast performances shown for different overarching modeling themes. A. Median of the scores of models which belong to four different themes shown for Weekly incidental COVID-19 Cases forecasts over time. B. Median of the scores of models which belong to four different themes shown for cumulative death counts forecasts over time.

## Conclusions and Discussions

We developed a scoring framework for the forecasts of COVID-19 epidemiological models using forecast and observed data available at COVID-19 Forecast Hub. Scores can be used to evaluate past performances of all models. Additionally, quantitatively evaluating model forecasts enable score-weighted ensemble model forecasting. We provide a systematic review of available model types which show what type of modeling efforts are more successful in forecasting the COVID-19 epidemiology in the US. Our results suggest that models are unable to capture abrupt changes in COVID-19 epidemiology (e.g. the first two weeks of July and the first two weeks of November in US). Death count forecasts so far have been more accurate than weekly case counts. That could be because incidental case count curves are much higher than death counts and non-monotonic in behavior and may have varying reporting practices across health institutions, counties, and states. We refer the reader to www.covidtracking.com/data for an overview of such variabilities.

Our work has some notable limitations. We only consider models on the COVID-19 forecast hub and this may inadertently lend to selection bias of groups willing to format their model output according to required metrics and upload to the hub. Currently, our scoring framework considers only the US national data and scores on the individual US state model forecasts can be made available. We have also only considered weekly incidental case numbers and number of cumulative deaths, which does not take into account modeling efforts that predict hospital capacity and utilization. Additionally, our ensemble model formation may be optimized. When reporting average forward score for a model, we give equal weights to forecasts made earlier in time to the more recent forecasts. The score-weighted ensemble forecasts might have performed better, if we had focused on the recent forecasts instead of the entire set of longitudinal data pertaining to the pandemic.

This work supplements the COVID-19 Forecast Hub effort by taking the modeler provided probability distributions and computing the score for each week the research groups update their forecasts. This can be implemented quickly, but does not standardize how the model uncertainty is computed. This, in particular, can be important if the model is a mechanistic one with multiple parameters. In such a case, the performance of the model should depend on the mechanisms included, the priors on the parameters in the model, and the chosen likelihood function for the noise on the data. Separating these effects might be useful in informing which mechanisms, priors, and noise models are important in obtaining accurate forecasts.

A more ambitious scoring framework would be to perform a Bayesian scoring analysis on all models in-house. This would be made easier if model codes were to adhere to a universal standard. The universal standard idea has already been implemented by the FDA for results of clinical trials using Biocompute Objects (13), which standardize the data processing pipeline. Universal model scoring would be optional but groups would be incentivized to obtain a score in order for their model to be considered as part of an ensemble. The standard could also be made less invasive by using the concept of the Unit test, which is an executable code snippet that verifies the intended behavior of a program. This is widely used in complex software projects. Google alone executes 4.2 million unit tests daily to verify the behavior of its core products. The analogue would be code snippets for computing the model score.

SciUnit (http://sciunit.io) is a Python package for constructing, executing, and reviewing the outcomes of model validation tests (14). The core design principle is the programming adage “separate the implementation from the interface”. Tests are not required to know anything about how a model works; instead, models expose functions that a test calls through an interface called a capability. The implementation of capabilities is left to downstream libraries, where SciUnit underlies libraries for a variety of projects, including The Human Brain Project (1000+ investigators) and OpenWorm (80 investigators across 17 countries). We could integrate public epidemiological models with SciUnit to rapidly, programmatically, and objectively assess the inferential and predictive performance of competing models across a number of relevant public health dimensions.

## Data Availability

All data and computer code used is publicly available

https://github.com/ONYLAB/Scoreboard

## Data and Code Availability

The code for the scoreboard is available at github.com/ONYLAB/Scoreboard.Our scores are updated every Sunday evening and available at www.covidforeca.st.

## ACKNOWLEDGEMENTS

MRM was supported in part by an appointment to the Research Participation Program at OBE/CBER, U.S. Food and Drug Administration, administered by the Oak Ridge Institute for Science and Education through an interagency agreement between the U.S. Department of Energy and FDA. CCC is supported by the Intramural Research Program of the NIH/NIDDK. The authors received no financial support from any source, and there is no conflict of interest. This article reflects the views of the authors and should not be construed to represent FDA’s views or policies.

## Supplementary 1: Supplementary Figures for “A Quantitative Evaluation of COVID-19 Epidemiological Models”

**Fig. S1.**
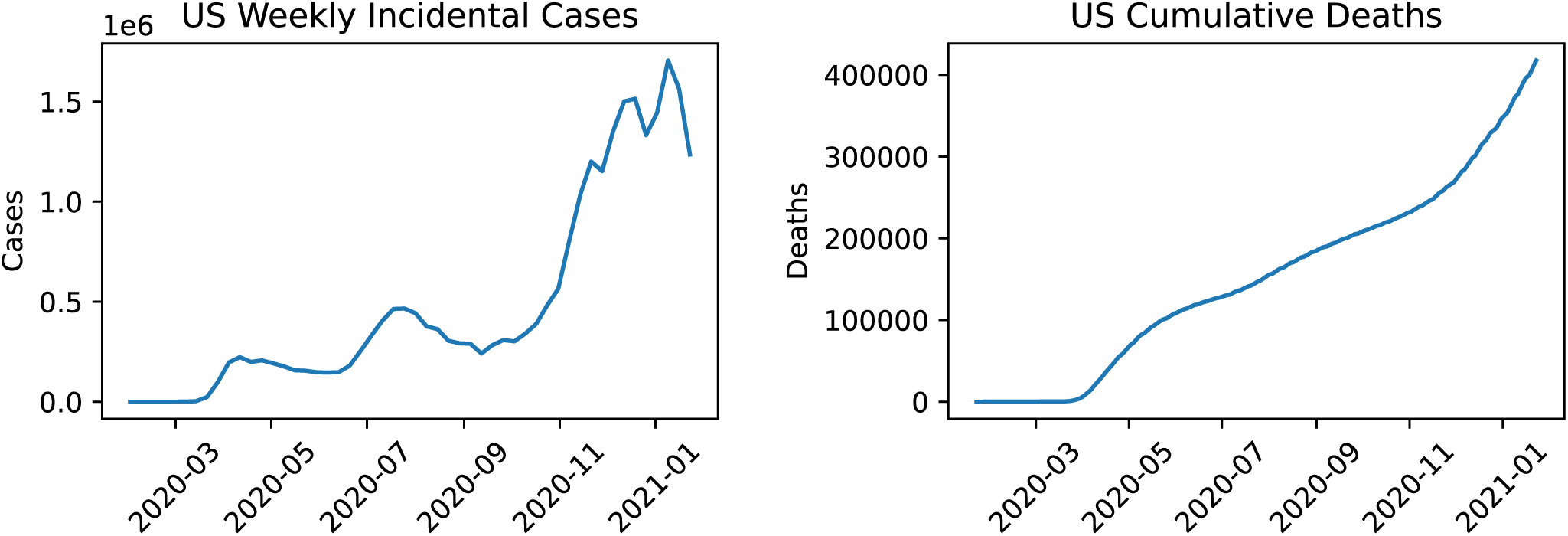
COVID-19 in the US

**Fig. S2.**
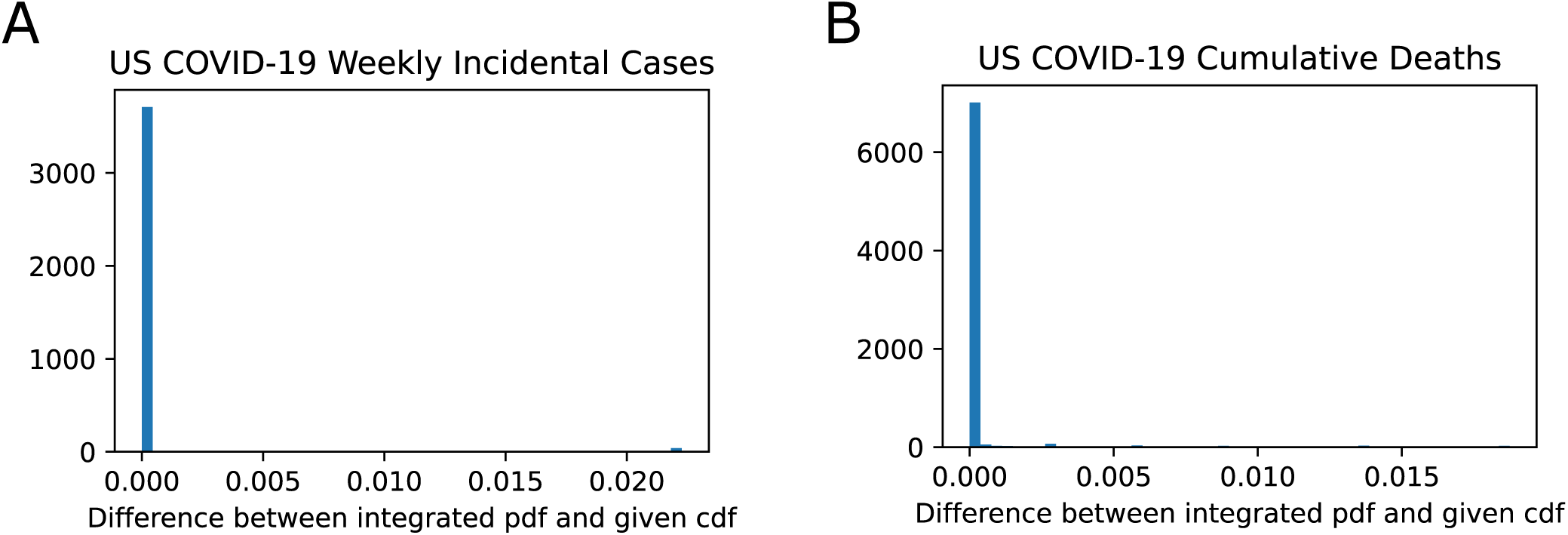
Overall quality of CDF to PDF Conversion.

**Fig. S3.**
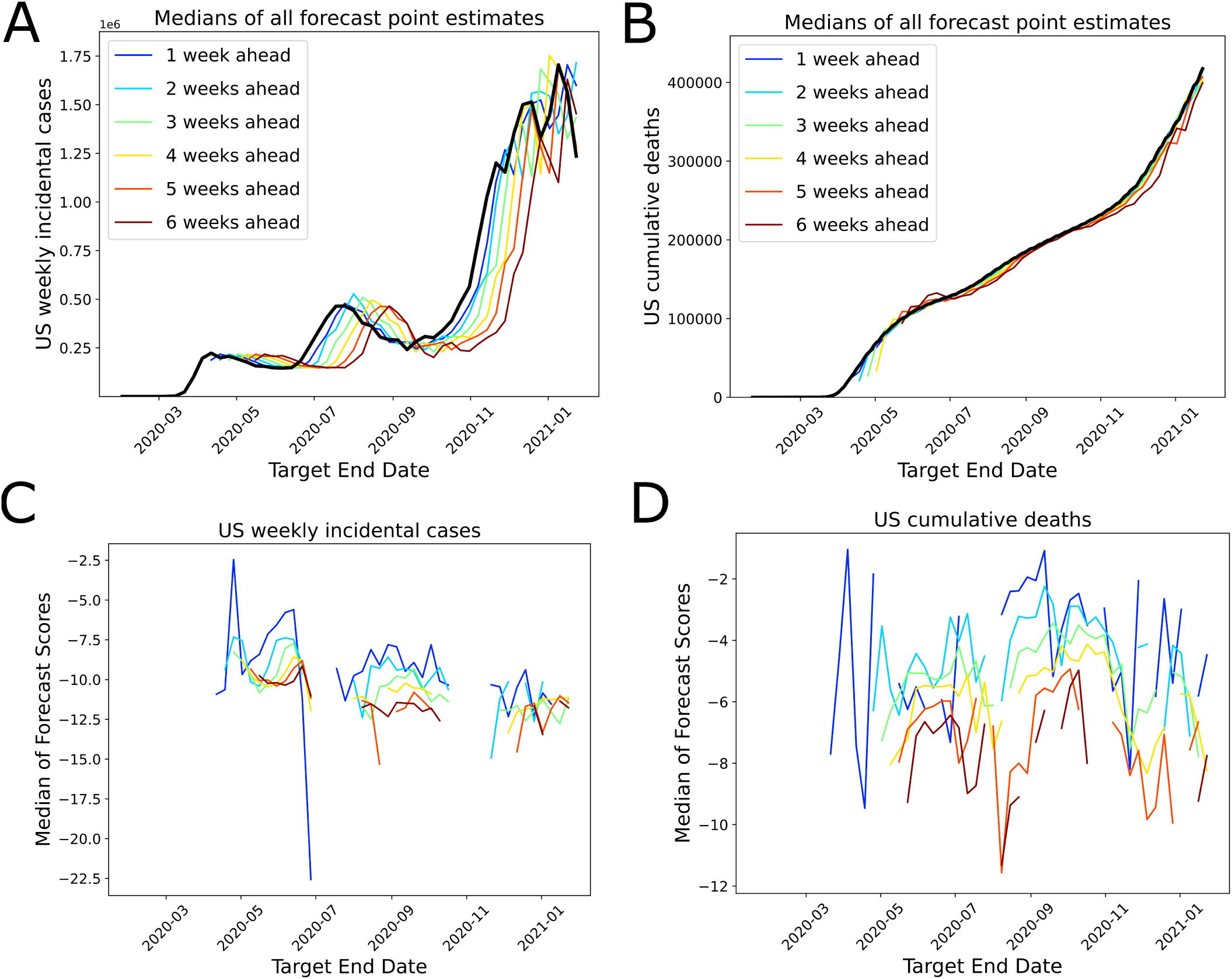
Model forecast performances over time. A. Black solid curve represents the observed US weekly incidental case counts. Other curves represent the median of the forecasts for the target end date made from 1 to 6-weeks prior to the target end dates. B. Black solid curve represents the observed US cumulative death counts. Other curves represent the median of the forecasts for the target end date made from 1 to 6-weeks prior to the target end dates. C. Curves represent the median of the forecast scores colored based on their forecasting horizon (1-week prior to 6-weeks prior color-matching to sub-panel A). D. Curves represent the median of the forecast scores colored based on their forecasting horizon (1 to 6-weeks prior, with colors matching to sub-panel B). Discontinuities in the score plots imply that the median value of the scores for a particular time-horizon is −∞, demonstrating the poor performance especially on the inflection points of the epidemiological curves.

**Fig. S4.**
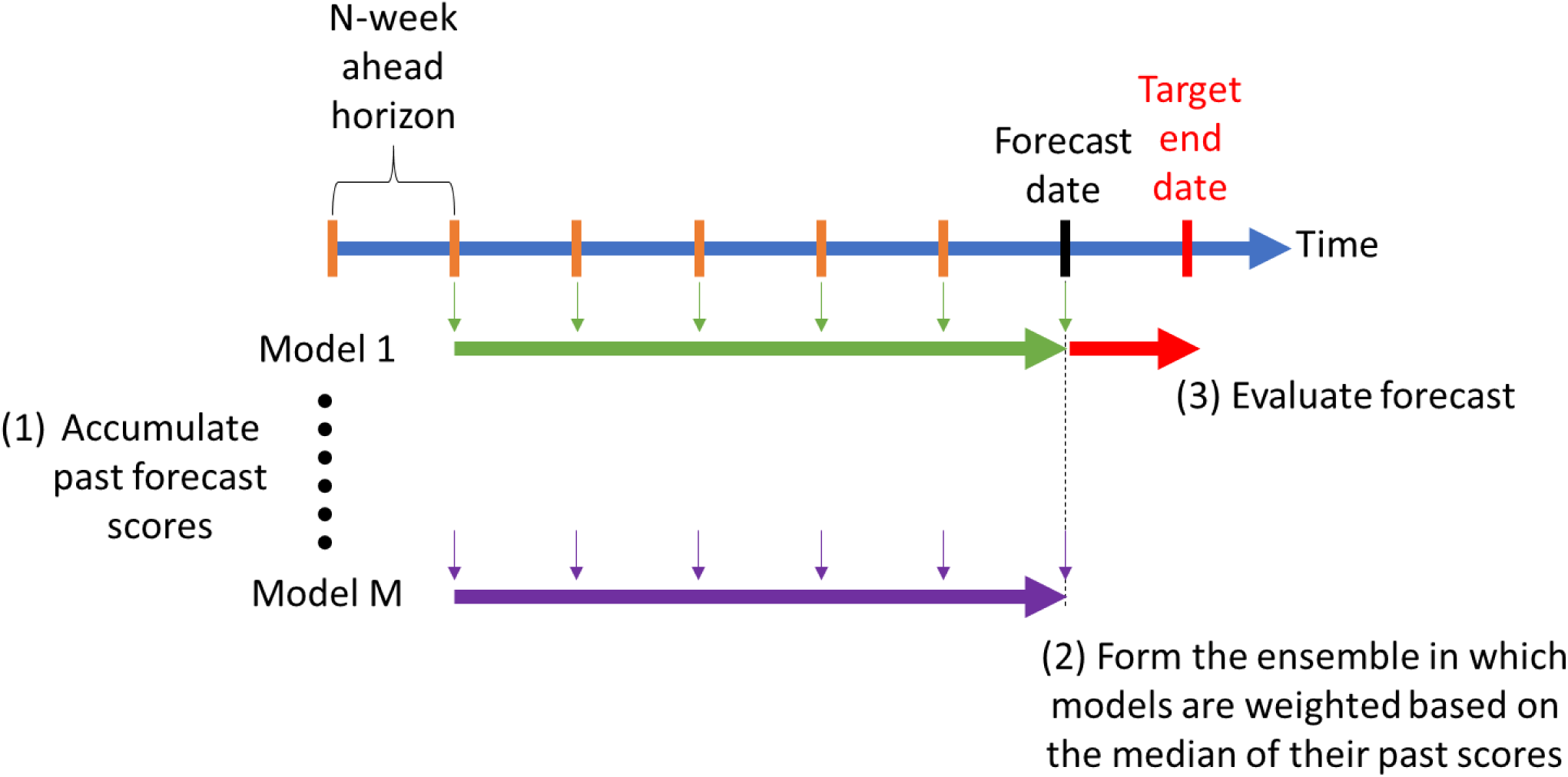
Ensemble forecast formation.

**Fig. S5.**
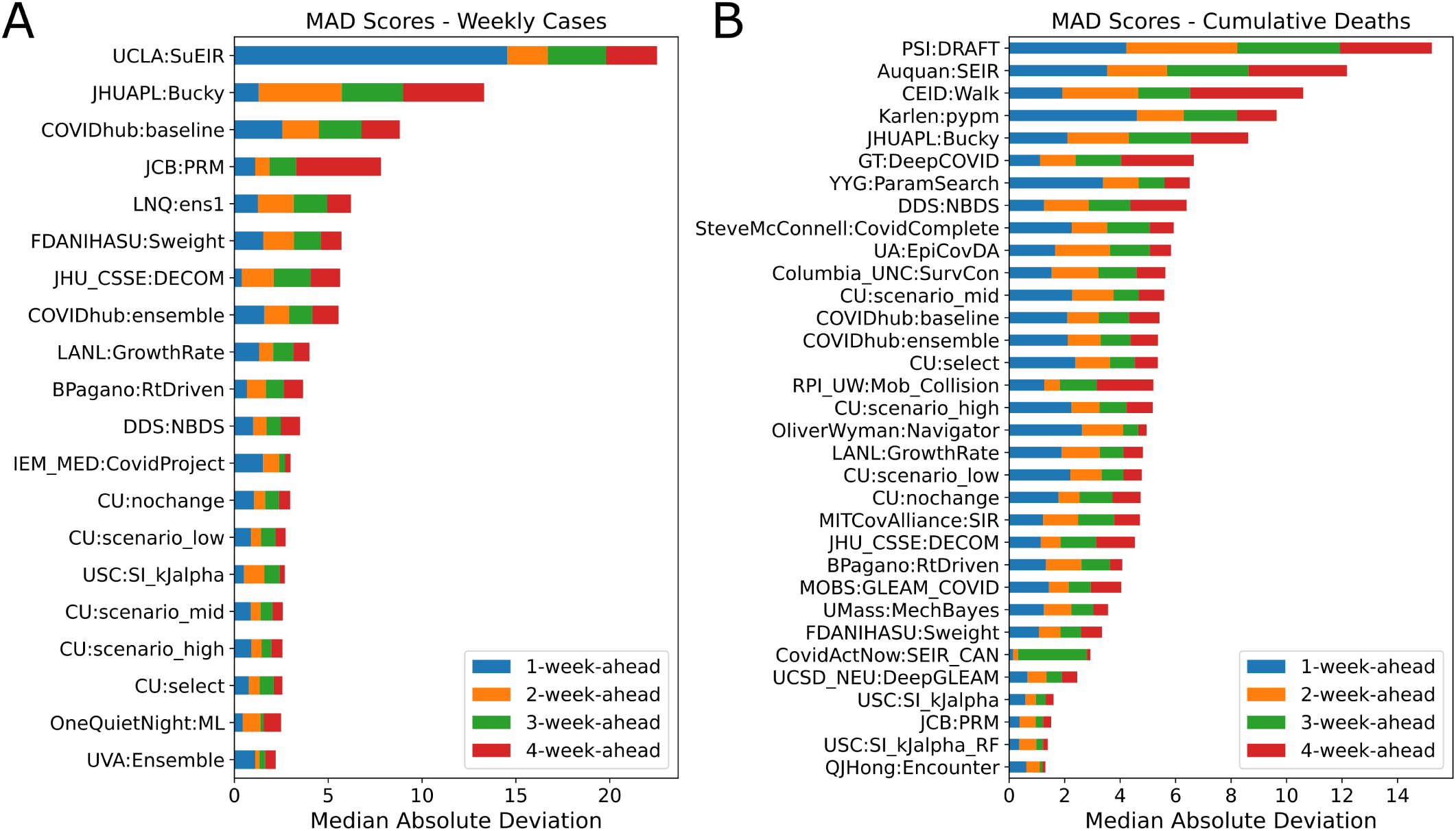
Median absolute deviation (MAD) as a score variability measure for the models. A. MAD for weekly incidental case forecasts over 1-4-week-ahead forecasting horizon. B. MAD for cumulative death count forecasts over 1-4-week-ahead forecasting horizon. FDANIHASU model is the score-weighted ensemble presented in this work. Note: Models that have at least one *∞* as their MAD in 1-to 4-week-ahead forecasts are not shown in these plots.

**Fig. S6.**
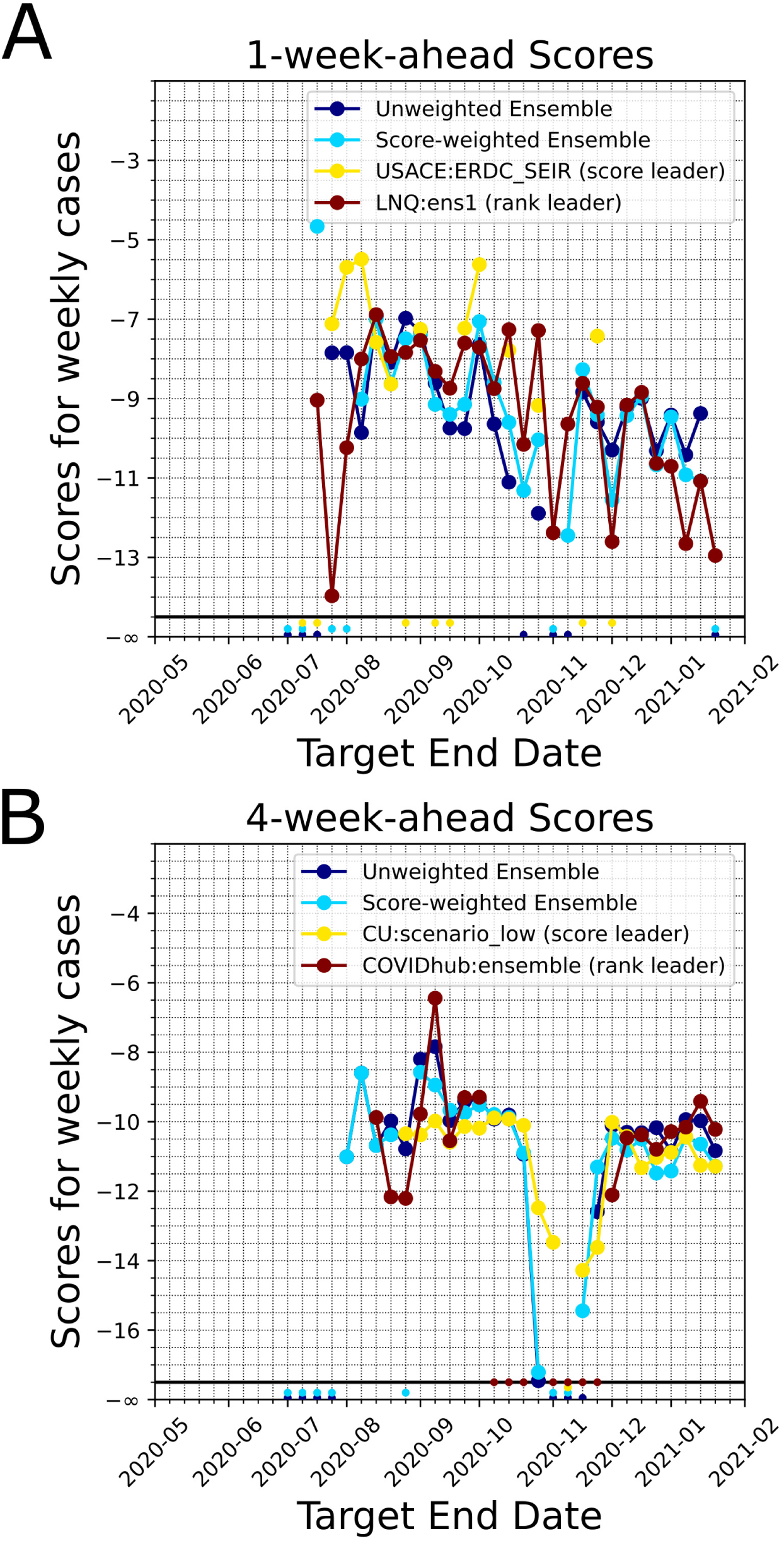
Comparison of scores of unweighted and score-weighted ensemble models for the weekly incidental case counts. For comparison purposes we plot the leading model’s score (based on past performance as of the last target end date, by ranking and by median scores). Model forecasts that do not encompass the ground truth *G* would have a score −∞ and this scenario is shown at the bottom of the figure panels. A. 1-week-ahead scores. B. 4-week-ahead scores.

**Fig. S7.**
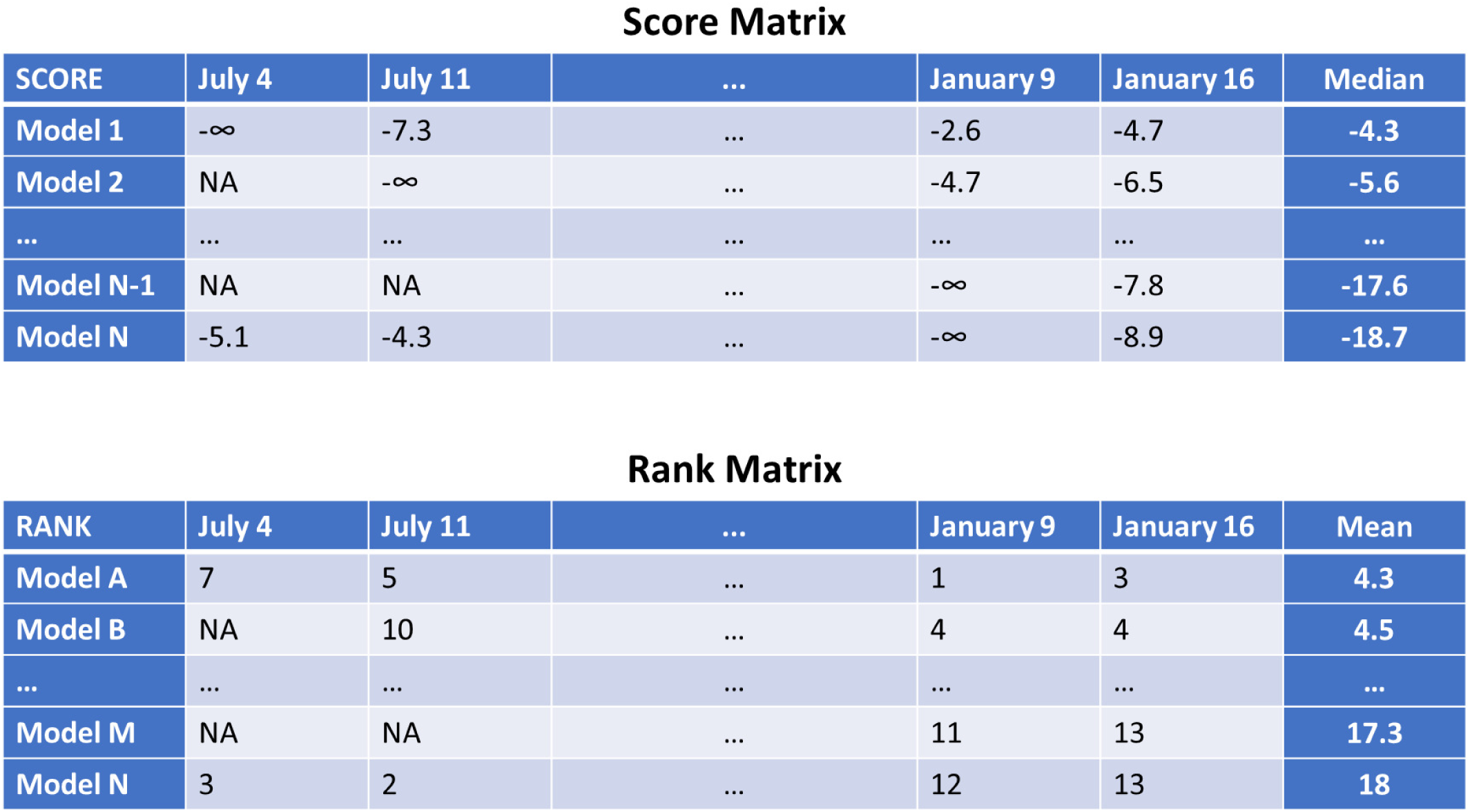
Example score and rank matrices for n-Week forecast horizon. Cells in the matrices with NA represent the absence of a forecast for that date. There are 12 categories and each category has its own leader board: 2 for targets (weekly cases, cum deaths) and 6 for time horizons (1-week-ahead,.., 6-week-ahead). We have two conditions in place when forming the leader boards using these matrices. First, the leader boards consider the time frame July 4, 2020 onward. Second, in the leader boards, we do not include models with number of forecasts less than 50% of the number of possible weeks in the time frame.

